# COVID-19 Intensive Care Admissions Have Twice the Corrected Mortality of non-COVID-19 Viral Pneumonia

**DOI:** 10.1101/2020.07.23.20161059

**Authors:** Dan V. Nicolau, Alexander Hasson

## Abstract

Studying the *ICNARC Case* Mix *Programme Database* has yielded results showing intensive care admissions by those infected with COVID-19 have twice the corrected mortality of patients presenting with non-COVID-19 viral pneumonia. A basis of an APACHE-II-like score denoted as “BASCA” is also outlined in this study.

---

The COVID-19 pandemic is placing unprecedented demands on healthcare systems worldwide and on intensive care units (ICUs). While most patients experience mild, self-limiting disease, a substantial proportion require hospitalisation, with the majority of these needing respiratory support and many going on to ICU admission^1^. Since COVID-19 appears to be a distinct clinical entity^2^, it is crucial to understand expected clinical outcomes for these patients, particularly in relation to those with (non-COVID-19) viral pneumonia, whose management is well studied^3^.

We studied data in the *ICNARC Case Mix Programme Database*^4^ up to the 4^th^ April, 2020. Adequate data was available for 2249 patients admitted to ICUs in England, Wales and Northern Ireland with confirmed COVID-19 either at or after admission to ICU. Of these, 346 (15%) had died, 344 (15%) were discharged alive from critical care and 1559 (65%) patients were last reported as still being in critical care.

We compared the admission characteristics of these patients with those with non-COVID-19 viral pneumonia admitted to ICUs in the UK between 2017-2019^4^. The mean age at admission was similar (60.1 vs 58.1 years), while COVID-19 patients were more likely to be male (73% vs 54%). ICU COVID-19 patients’ mean BMI was smaller (∼22.5) than non-COVID-19 viral pneumonia patients, whose BMI reflected that of the general population when matched for age and gender (∼27.5). COVID-19 patients were healthier across the board prior to admission, with less requiring living assistance (7.2% vs 26.5%), having cardiovascular disease (0.3% vs 1.3%), respiratory illness (1.1% vs 4.9%, liver disease (0.2% vs 0.9%), metastasis (0.4% vs 1.2%), haematological malignancy (1.1% vs 4.4%), immunocompromise (2.3% vs 8.5%) and mean APACHE-II score (5.1 vs 6.2) at admission.

We also compared the outcomes for the two cohorts. 62.9% of COVID-19 ICU patients required ventilation within the first 24 hours, compared with 42.6% for viral pneumonia patients. Combined mortality for COVID-19 was roughly twice as high (50.1% vs 22.4%). *Remarkably, all else being equal, the case mortality of COVID-19 ICU admissions is twice that of non-COVID-19 viral pneumonia (Figure 1), except for the BMI>30 cohort (3*.*1 times higher)*.

**Figure 1.**
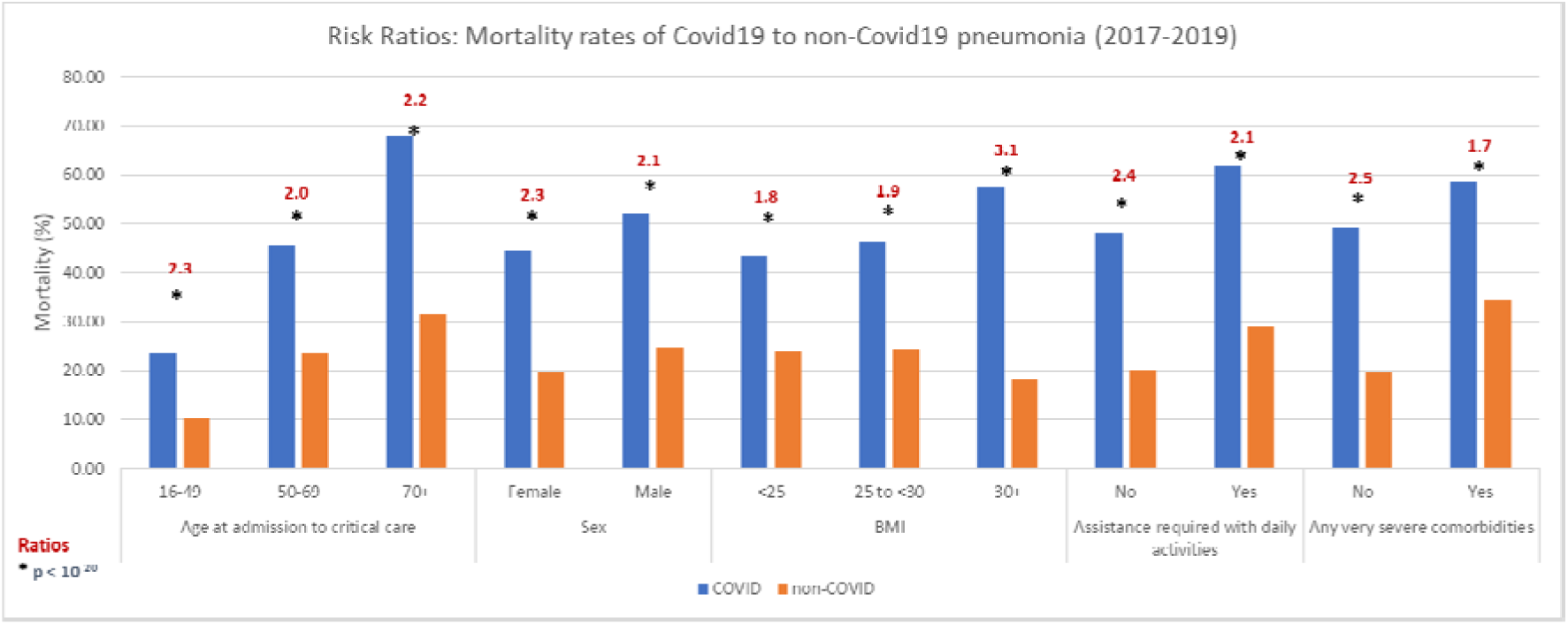
Case mortality for ICU admissions with COVID-19 (blue) vs. non-COVID-19 viral pneumonia (orange), stratified by characteristics at ICU admission. Mean RR between COVID-19 vs non-COVID-19 pneumonia ICU admissions = 2.2.

Limitations of our analysis include discharge status, since many of the patients in the database were still in ICU, as well as coarse stratification by age, BMI and existing comorbidities, which we will address in future work, once more data is available.

Nonetheless, taken together, these early data suggest that age, gender, severe comorbidities and BMI contribute to mortality in COVID-19 ICU patients, perhaps in a roughly additive way. As a rule of thumb, consistently with our data here, we suggest calculating a cumulative score from 1-8, using the mnemonic “BASCA”, as follows: BMI (<25=0; 25-30=1; 30+=2) + Age (<49=1; 50-69=2; 70+=3) + Sex (Female=0; Male=1) + Comorbidities (No=0; Yes=1) + Assistance (No=0; Yes=1). Multiplying the result by 10 gives an estimate of the percentage mortality for a given patient at ICU admission. Thus, a male patient (+1) of 55 years (+2) with existing comorbidities (+1) and a BMI of 27 (+1) would have an expected mortality of approximately 50%. Validation of this score will be undertaken as more data becomes available.

## Data Availability

Availability of data and materials: raw data is available from; the ICNARC database and our processed data on request from the authors.

## Declarations

Ethics approval and consent to participate: none required. Consent for publication: not applicable. Availability of data and materials: raw data is available from; the ICNARC database and our processed data on request from the authors. Competing interests: none. Funding: DVN is supported by a Future Fellowship of the ARC. Authors’ contributions: DVN conceived the study, analysed the data and wrote the Letter; AH collected and analysed the data. Acknowledgements: none.

